# Leveraging an Online Dashboard to Inform on Infectious Disease Surveillance: A case Study of COVID-19 in Kenya

**DOI:** 10.1101/2024.09.14.24313681

**Authors:** Mike J. Mwanga, Laura M. Guzman-Rincon, Leonard Kingwara, Don B. Odhiambo, Henry Gathuri, Arnold Lambisia, John Morobe Mwita, Edidah Moraa, Bernadette Kutima, John Gitonga, Daisy Mugo, Charles N. Agoti, James Nyagwange, George Warimwe, Isabella Oyier, D. James Nokes, Ambrose Agweyu, E Wangeci Kagucia, Anthony Etyang, John Kiiru, George Githinji

**Affiliations:** KEMRI-Wellcome Trust Research Program, Kilifi, Kenya; School of Life Sciences and Zeeman Institute for Systems Biology and Infectious Disease Epidemiology Research (SBIDER), University of Warwick, Coventry, UK; Ministry of Health, Nairobi, Kenya; Kenya National Public Health Institute, Nairobi, Kenya; London School of Hygiene and Tropical Medicine, London, United Kingdom; Department of Biochemistry and Biotechnology, Pwani University, Kilifi, Kenya

**Keywords:** COVID-19, SARS-CoV-2, genomics, dashboard, dash, python, Kenya

## Abstract

A multi-pronged approach to combating the COVID-19 pandemic in Kenya resulted in the formation of multidisciplinary research initiatives including genomic sequencing, syndromic surveillance, sero-surveillance, vaccination, and mathematical modelling. These initiatives generated an overwhelming amount of data that posed a challenge to researchers and public health officials, to effectively manage, analyse and promptly interpret for immediate pandemic response. As a result, there was demand for a platform to collate and integrate these datasets with interpretable findings to aid in pandemic management. In response, we developed a web-based dashboard, and integrated multidisciplinary datasets collected by the Ministry of Health-Kenya (MoH-K) and other research organizations, to support surveillance and monitoring of COVID-19 in Kenya. The developed dashboard combines genomics, epidemiological, seroprevalence, modelling, vaccination, syndromic and phylogenetic data and provides real-time updates to the public and health sector experts. The dashboard provides temporal trends of reported COVID-19 cases, fatalities, variants, and vaccination, in addition to summary reports from multiple cross-sectional seroprevalence studies and ongoing facility-based inpatient syndromic surveillance from 15 health facilities across Kenya. This is the first detailed interactive dashboard in Kenya that combines multiple datasets from a disease outbreak to provide valuable insights to researchers, health policy makers, the media and public not only during pandemic but also during routine surveillance. This resource is a model for digital platform for infectious disease surveillance and for informing public health planning and intervention.

**Dashboard Link:** https://kcd.kemri-wellcome.org/

## Introduction

The emergence of the COVID-19 pandemic triggered unprecedented challenges to governments and the public health sector, especially the need for high-quality, locally relevant and timely data, important for informed policy and decision-making (1). Various partnerships had to be formed among public and private institutions including local governments to collect and collate relevant and urgently needed data for pandemic management (2). In Kenya, multidisciplinary research initiatives including genomic sequencing, syndromic surveillance, serosurveillance, vaccination and mathematical modelling for predictions were established. These initiatives resulted in ever-increasing data ranging from virus genetic makeup to patterns of disease spread and population immunity. The wealth of data from multiple disciplines and spanning different time points became overwhelming not only to scientists but also to policy makers. This, coupled with the fact that there were limited channels to communicate and report findings from these data to the public necessitated the need for streamlined approaches to cohesively integrate and present multidisciplinary data. This led to an increased demand for efforts to implement innovative technologies and tools to integrate multidisciplinary data and facilitate evidence-based decision making for effective disease control policies especially during disease outbreaks. Given these challenges, leveraging information technology solutions such as dashboards is indispensable to collecting, managing, analysing, and displaying data from various sources and assisting in decision-makers in making informed decisions and appropriate interventions.

Development of integrated online dashboards can significantly improve infectious disease surveillance by bringing together multiple data sources such as clinical, genomic, epidemiological, and other complimentary metadata and identify trends which in turn can provide insights for effective disease intervention and control (3). Dashboards have been previously used to communicate, manage, and guide responses to outbreaks and epidemics by providing up-to-date, comprehensive, and interactive visualizations of public health data. For instance, the Somalia Polio outbreak of 2013-2014 (4) utilized a dashboard as a utility for monitoring and rapid responses. The ongoing COVID-19 pandemic has provided a rich environment within which dashboards have proliferated, such as the John Hopkins University dashboard (5), World Health Organization (WHO) (6), and Africa Centre for Disease Control (7). Such dashboards were effective in providing high level of global trends used in tracking the progress of the COVID-19 pandemic and facilitating information sharing among governments, collaborators and healthcare providers.

We developed a web-based dashboard, and integrated multidisciplinary datasets collected by the Ministry of Health – Kenya (MoH-K) and other research organizations, to support COVID-19 disease surveillance aimed at contributing pandemic management and timely decision-making process. We utilized COVID-19 datasets from multiple surveillance data streams that were established to enable tracking and monitoring of COVID-19 epidemic. In addition to the SARS-CoV-2 demographic, spatial, and temporal trends, we incorporated growth-rate estimations for recently identified lineages using a Bayesian hierarchical model (8). The model estimates growth rate of selected SARS-CoV-2 lineages and serves as a warning system on the potential increase in frequency of the variant in Kenya (see Supplementary file).

## Data and Data Sources

### Clinical and epidemiological data

The COVID-19 clinical, and epidemiological data were collected through a network of sites that includes both public and private health facilities across the country. Sample associated metadata are captured using standard case investigation forms provided by MoH-K. Research organizations and health facilities are mandated to report testing results and vaccination reports to MoH-K (Figure 1). These reports are also accompanied by patient demographic and clinical data (date of birth, county of residence, travel history, age, and medical history, such as other comorbidity infections) which are then collated to make a country-wide linelist. These data were summarized and visualized within the home and county panels.

**Figure 1:**
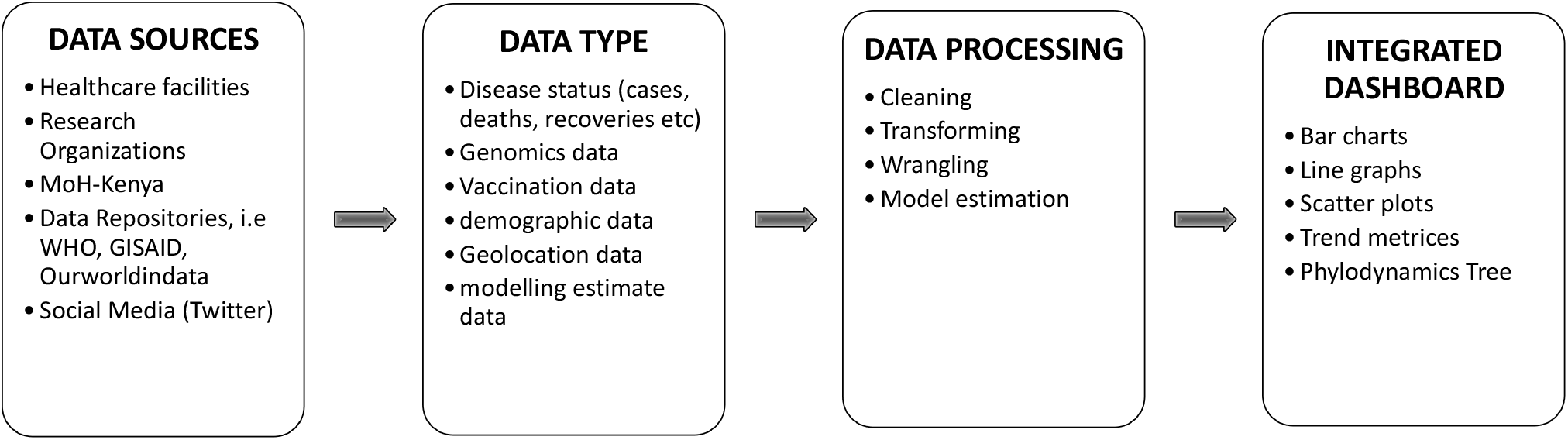
Schematic presentation of data workflow and processing stages and integration to the dashboard.

### Vaccination data

Data related to vaccinations, including demographic information, dosage, and county-level vaccination information, were obtained from the MoH-K online resources. These data is displayed under the vaccination panel.

### Genomic and modelling data

Dedicated sequencing facilities performed weekly sequencing of SARS-CoV-2 positive samples and in turn submitted the sequence data to public repositories including GISAID and GenBank. The submitted sequences included associated sequence metadata such as location of sample collection, age of patient, date of sample collection and submission, variant genotype, and submitting lab. These data were leveraged to enable understanding of countrywide virus diversity and spread; data which is visualized under the variants tab. The sequence data was also used to infer evolutionary relationships using the Nextstrain genomics platform (9). Nextstrain hosts a bioinformatics pipeline for phylodynamic analysis that integrates genomics sequence data with other data types such as geographic, epidemiologic, and host species. We utilized this interactive visualization platform to track evolution and spread of SARS-CoV-2 virus in Kenya by integrating the results in phylogenetics tab. Additionally, the genomic data on circulating variants and lineages was used in estimating variant growth rates using a novel Bayesian growth rate model (8). The choice of lineages for the growth-rate analysis is contingent on the weekly sequencing results and publicly available sequences from other sequencing centres in Kenya. The growth model takes newly identified and emerging SARS-CoV-2 lineages in Kenya and compares their frequency and growth rates to those observed in other countries from Africa, North and South America, Europe, and Oceania. Text files containing the weekly frequency of summarized SARS-CoV-2 lineages in Kenya and the stated regions are used as input into the model which then estimates lineage growth rates. Lineages showing consistent increase in abundance for at least two consecutive weeks in Kenya and/or any of the regions/country will be flagged as a cautionary signal in red. We used this to include an early warning system which can effectively be used to signal rising cases of lineages and be used to facilitate decision making at national and sub-national levels.

### Serosurveillance data

Other surveillance studies, such as seroprevalence studies (10–12) provided serology datasets that informed the level of COVID-19 infection within the population in the early phases of the pandemic in different study populations.

### Adult syndromic surveillance data

In addition, countrywide datasets from the Adult Syndromic Surveillance work were added to provide a disease situational awareness which is important in national pandemic preparedness plans. This is achieved through a hospital-based surveillance program set up as part of national response to COVID-19 in Kenya. The initiative collects daily clinical data from adults admitted in medical wards in at least 15 health facilities in Central, Coast and Western regions of Kenya.

## Kenya COVID-19 Dashboard Architecture

### Framework and data integration

The Kenya COVID-19 dashboard was prototyped using the Dash Python framework, an open-source and freely accessible web framework written on top of Flask, Plotly and React frameworks. The source code is written in Python (v3.10) language and the dashboard is hosted on an Apache (v2.4) web server running on a Linux environment (Ubuntu v22.04).

Tabular spreadsheets served as input data files that were continuously updated weekly and monthly, Supplementary table 1. In the backend, Pandas and NumPy python libraries were used in parsing and processing of the data files. The front end is implemented using the Plotly Dash package (Plotly Technologies Inc. 2015, https://plotly.com/python/) to structure and display pages, handling user callbacks and visualize the data (figure 2).

**Figure 2:**
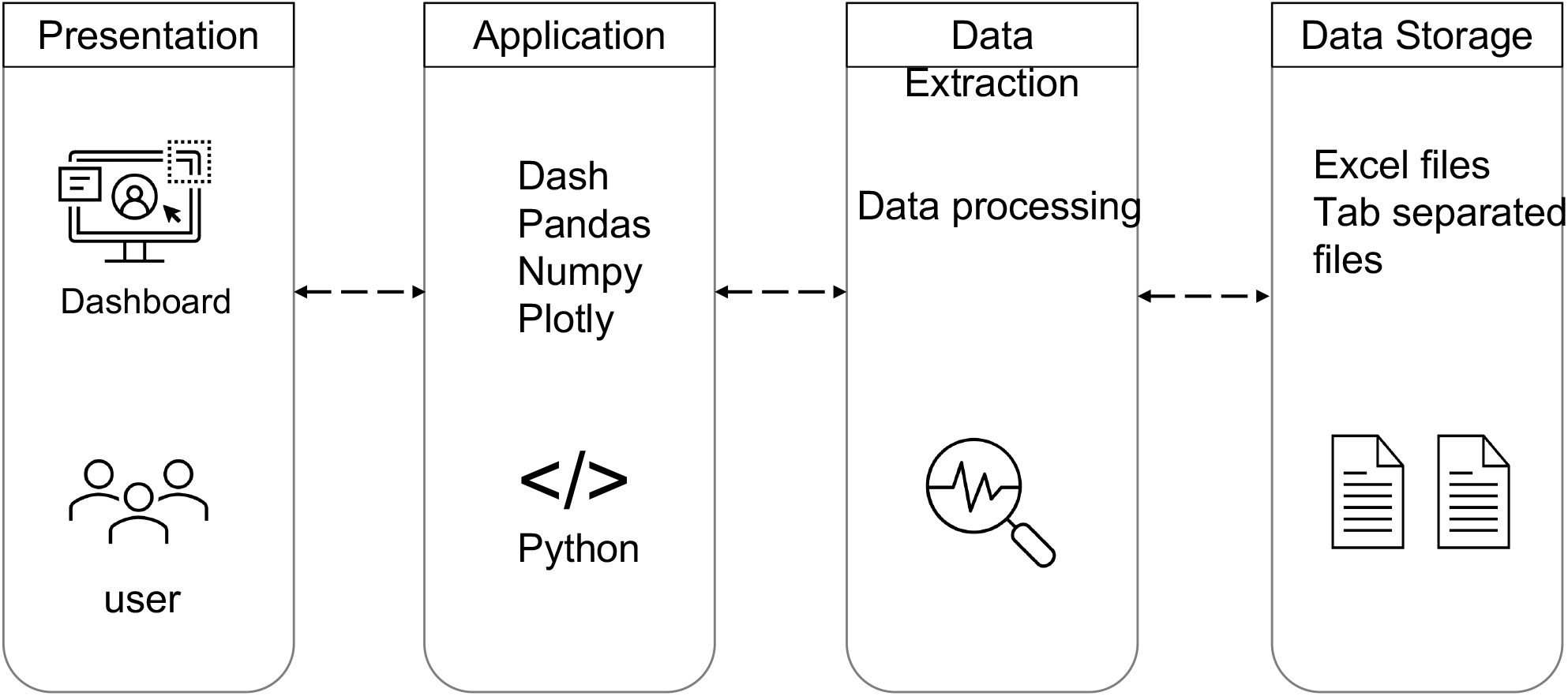
Illustration of the Kenya COVID-19 dashboard architecture from backend to frontend. When users select on components on the dashboard, python libraries are utilized to query and extract the dataset from stored files, generate visuals that are then displayed on the users’ screen.

Prior to being visualized in the dashboard, the datasets undergo rigorous preprocessing steps to clean, transform, and validate the raw data and ensure accuracy and completeness. Cleaning process involved identifying and correcting wrong date formats, value mislabelling (i.e count and sub-county names and hospital names). Aggregated summaries for different patient age categories, counties, and temporal were performed on the linelist datasets. Similarly obtained serosurveillance and syndromic surveillance datasets were summarized and visualized by region, gender, age categories and study populations.

Estimates from the Bayesian growth rate analysis (8) are calculated separately using R (version 4.4.2) and we used this to include an early warning system that was relevant in informing increase in cases. The resulting data is rendered using plotly graph package in the dashboard. Any newly identified SARS-CoV-2 lineages observed in Kenya are updated in the model dataset and subsequently an update of the growth rate estimation results.

The phylogeny tab uses Nextstrain genomics pipeline (9) to generate phylogenetic relationships for SARS-CoV-2 lineages identified on samples collected within Kenya. The generated analyses configuration files (https://github.com/veclab/ncov-kenya) are shared through the Nextstrain community URLs (https://nextstrain.org/community) and mapped onto the dashboard. The use of open-source tools and the availability of the associated COVID-19 dataset allowed rapid customization of the platform. The source code is placed in a GitHub repository and can be accessed online (https://github.com/mikemwanga/Kenya-COVID-19-Dashboard). The dashboard is maintained by KWTRP and can be accessed via this link: https://kcd.kemri-wellcome.org/.

### Dashboard Pages

The beta version of the multi-page dashboard is currently running and displays data reports on COVID-19 surveillance and monitoring in Kenya. The dashboard is tabbed with multiple pages, i.e., home, county, seroprevalence, vaccination, syndromic-surveillance, variants, and phylogeny.

The home page provides a summary of the most recent data on the total number of cases, and tests carried out, SARS-CoV-2 positivity, deaths, and recoveries reported since the beginning of the pandemic. This page also displays the number of cases, deaths, tests done, and positivity in a span of 24-hour period as reported by MOH-K. Additionally, graphical displays indicating counties with reported cases in the last 24 hours, a breakdown of the counties with reported cases the countrywide epidemiological curve of cases and fatalities, and cases and deaths in different age groups and gender categories are described.

The county page is designed to allow visualization of COVID-19 epidemiology at the sub-national level. Users interactively filter by county of interest and view the selected county’s total cases, total fatalities, and overall positivity. Additionally, graphs showing temporal daily trends in cases, and cumulative cases are displayed. The vaccination page overviews reported metrics of the number of persons vaccinated countrywide and percentage per county.

The seroprevalence page uses data from the serological studies conducted among different populations since the beginning of the pandemic. The users can interactively compare the change in seroprevalence between populations and on demography such as age, gender, and geographical location.

The Adult syndromic surveillance section provides a detailed summary of information captured from patient record files obtained from collaborating health facilities across the country. This section includes detailed information on patient demographics categorized by region and admission period, as well as symptoms and conducted tests during admission. Additionally, the section encompasses documentation of critical priority indicators and measures, such as COVID-19 vaccination status, HIV status, and recording of vital signs like temperature, blood pressure, and respiration rate.

The variant trends page seeks to understand temporal trends and variations of circulating SARS-CoV-2 variants and lineages. Here, temporal prevalence data for the most recent circulating SARS-COV-2 lineages across the country is also viewed. Furthermore, results of the growth rate model for warnings on lineages of concern are also displayed in this page. The phylogeny tab allows phylogenetic tree visualization of SARS-CoV-2 lineages using Nextstrain bioinformatics pipeline for phylodynamic analysis and interactive visualization. The nextstrain platform is used to analyse and compare lineages observed locally against a background dataset of local and global lineages. The availability of county metadata enables an understanding of lineage diversity at both national and subnational levels.

### Significance

The Kenya COVID-19 dashboard was completed and deployed by August 2023. By the time of writing, the platform reported over 344,000 COVID-19 cases, 5689 deaths and an overall positivity of 8.6% by December 2023. Cases were most reported in individuals between ages 21-40 in both male and female, while high fatalities was experienced in adults between 61-70 years of age. COVID-19 cases were most prevalent in Nairobi (41.3%), Kiambu (6.27%), Mombasa (5.38%), Nakuru (5.2%) and Uasin Gishu (3.21%) counties. The rate of infection was highest in Nairobi County where at least 3.18% of the population were infected, while the in the rest, >1% of the population were infected.

Vaccination data from MoH-K shows that only 63% of the over 29 million received doses were administered and that 37.2% of the Kenyan population are fully vaccinated. Vaccination with first and second doses was high in persons of ages between 18-29 and 30-39 years while most of the partially vaccinated persons were of ages between 15-18 years. At county levels, over 50% of Nairobi (56.3%), Nyeri (54.3%), Siaya (52.2%) and Kisumu (51.2%) residents had been fully vaccinated. In contrast the least vaccinated populations were from Mandera (10.6%), Wajir (11.2%), Tana River (11.7%), Isiolo (13.6%), Marsabit (14.1%) and Garissa (14.4%) counties.

Sero-surveillance data shows significantly high (93.4%) seroprevalence by December 2022 as demonstrated by high anti-S IgG levels within the Health Demographic System Surveillance (HDSS) residents. The anti-S IgG levels were significantly lower (4.5%)_in the initial phases of the pandemic as observed in studies done on blood donors between December 202 to April 2021.

By end of December 2023, syndromic surveillance analysis was performed on 25,709 admitted in multiple health facilities from Wester, Central and Coastal regions. Documentation of respiratory rate, heart rate and blood pressure improved from April 2023 to December 2023 where 100% of all patients had these measures recorded. In contrast, less than 70% of the patients had their body temperature and heart rate recorded and less than 30% of patient’s had their height and weight recorded. There was variation in documentation of patient’s HIV status (36 to 70.1%) and COVID-19 (2.9% to 54.5%) vaccination between January to December 2023.

Towards the end of 2023, over 12,000 SARS-CoV-2 sequences from Kenya had been submitted to the GISAID platform. We summarised this data to provide and overview of temporal diversity of SARS-COV-2 variants and lineages. Generally, the dominance of non-variants of concern between March 2020 to January 2021, was replaced by co-circulation of alpha, beta, eta and delta variants of concern between January to November 2021. Since then, the omicron variant has dominated to December 2023. Diverse lineages of the omicron variant have co-circulated in 2023 with dominance of the FY.4.1-like and GE.1-like lineages.

## Discussion

Control and management of emerging infectious disease outbreaks require appropriate responses through optimal disease surveillance, monitoring, and timely outbreak detection which will inform among other things, the affected population, disease prevalence, and pathogen dynamics. Successful responses and outbreak management require the exploitation of all available data from a variety of disciplines to inform response in real-time and allow evidence-based decision-making. Continued use of conventional and traditional approaches have proved less effective in controlling such crisis hence the aspiration to integrate and utilize information technology systems such as dashboard in monitoring and tracking disease progression.

The Kenya COVID-19 dashboard provides a one-stop digital site to view up-to-date daily and long-term summaries of COVID-19 disease and can be used to develop intervention plans. We demonstrate the significance of integrating multiple datasets from a disease outbreak to support monitoring and enhance intervention measures. This is the first detailed interactive COVID-19 platform in Kenya that combines genomics, epidemiological, seroprevalence, modelling, vaccination, syndromic and phylogenetic data and provides real-time updates to the public and health sector experts. The dashboard provides simple and clear graphics to the users and allows interaction and customization of visuals. The dashboard makes complex genomics and epidemiological data accessible to a wider audience and can be used to inform public health policy and guide public health responses in Kenya. Making such visualization tools accessible to all partners and the public reduces time spent on other reports and routine data analysis. We demonstrate that it is possible to develop a ready-to-use and easy-to-setup online visualization dashboard using open-source tools that can serve as a platform for surveillance and monitoring of infectious diseases for outbreak response and interventions.

COVID-19 pandemic emphasized utilization of dashboards to track disease and be an integral part of public health disease surveillance and response efforts. During the unfolding of the pandemic, many countries struggled to make data publicly available and only few dashboards such as the Johns Hopkins University COVID Tracker became the to go for daily cases, deaths, tests among other metrices (5). Overtime, national and subnational dashboards have been developed to assist state agencies in managing the pandemic and expounding further to monitoring infectious pathogens. For instance, the United Kingdom Health Surveillance data dashboard was initially designed to report on COVID-19 pandemic but has further integrated other respiratory pathogens and seasonal influenza (13). The developed Kenya COVID-19 dashboard not only informs on COVID-19 disease trends but also on other COVID-19 related datasets such as variant diversity and seroprevalence. This signifies the importance of dashboards in providing key insights to researchers, health policy makers and public not only during pandemics but also during routine surveillance.

## Strengths and Limitations

The development and deployment of this dashboard uses free open-source tools and software and simple data techniques to achieve its aims. Python programme based free libraries such as pandas, NumPy, and plotly dash have been used to process, and render summary plots in this platform. This is a user-friendly and intuitive platform that is publicly and globally accessible with interactive features that allow users to select and compare data up to subnational levels. Furthermore, unlike most of other COVID-19 dashboard, integrating and displaying trends from multi-disciplinary sources namely case data, genomic, vaccination and seroprevalence allows conclusive understanding of disease outbreak.

On the downside, at the time of writing, there is reduced interest in COVID-19 disease by the target groups including public and policy makers; hence there is need to engage stakeholders and relevant authorities on significance of such platforms for future pandemic preparations. Additionally, the decline in COVID-19 prevalence has reduced data collection hence ‘slowness’ in updating the platform.

## Data Availability

Users who wish to access and use the data should send a request to the KEMRI Wellcome Trust Research Programme data governance committee, which can be contacted by emailing.

## Acknowledgment

We thank the Ministry of Health Kenya, and National Public Health Laboratories (Kenya), for sharing COVID-19 datasets. We thank members of the Pathogen Epidemiology and Omics group (PEO) and KEMRI Welcome Trust Research Programme for their contribution to this work.

## Code and Data Availability

The source code for the dashboard is placed in a GitHub repository and can be accessed online (https://github.com/mikemwanga/Kenya-COVID-19-Dashboard). Users who wish to access and use the data should send a request to the KEMRI Wellcome Trust Research Programme data governance committee, which can be contacted by emailing.

## References

1. Addai BW, Ngwa W. COVID-19 and cancer in Africa [Internet]. Vol. 371, Science.American Association for the Advancement of Science; 2021 [cited 2024 Feb 2]. p. 25–7. Available from: https://www.science.org/doi/10.1126/science.abd1016

2. Herman-Roloff A, Aman R, Samandari T, Kasera K, Emukule G, Amoth P, et al. Adapting Longstanding Public Health Collaborations between Government of Kenya and CDC Kenya in Response to the COVID-19 Pandemic, 2020-2021. Emerg Infect Dis [Internet]. 2022 Dec 1 [cited 2024 Feb 2];28(13):S159–67. Available from: https://www.nc.cdc.gov/eid/article/28/13/21-1550_article

3. Khodaveisi T, Dehdarirad H, Bouraghi H, Mohammadpour A, Sajadi F, Hosseiniravandi M. Characteristics and specifications of dashboards developed for the COVID-19 pandemic: a scoping review [Internet]. Journal of Public Health (Germany). Institute for Ionics; 2023 [cited 2024 Feb 2]. Available from: https://link.springer.com/article/10.1007/s10389-023-01838-z

4. Kamadjeu R, Gathenji C. Designing and implementing an electronic dashboard for disease outbreaks response - Case study of the 2013–2014 Somalia Polio outbreak response dashboard. Pan Afr Med J [Internet]. 2017 [cited 2023 Apr 13];27(Suppl 3). Available from: https://pubmed.ncbi.nlm.nih.gov/29296157/

5. Dong E, Ratcliff J, Goyea TD, Katz A, Lau R, Ng TK, et al. The Johns Hopkins University Center for Systems Science and Engineering COVID-19 Dashboard: data collection process, challenges faced, and lessons learned. Lancet Infect Dis [Internet]. 2022 Dec 1 [cited 2023 Apr 13];22(12):e370–6. Available from: https://pubmed.ncbi.nlm.nih.gov/36057267/

6. WHO. WHO Coronavirus (COVID-19) Dashboard | WHO Coronavirus (COVID-19) Dashboard With Vaccination Data [Internet]. 2022 [cited 2023 Apr 13]. Available from: https://covid19.who.int/

7. CDC. CDC. (2020). COVID-19 dashboard [Internet]. 2020 [cited 2023 Apr 13]. Available from: https://africacdc.org/covid-19/

8. Guzmán-Rincón LM, Hill EM, Dyson L, Tildesley MJ, Keeling MJ. Bayesian estimation of real-time epidemic growth rates using Gaussian processes: local dynamics of SARS-CoV-2 in England. J R Stat Soc Ser C Appl Stat [Internet]. 2023 Jun 27 [cited 2023 Aug 10]; Available from: 10.1093/jrsssc/qlad056

9. Hadfield J, Megill C, Bell SM, Huddleston J, Potter B, Callender C, et al. Nextstrain: real-time tracking of pathogen evolution. [cited 2023 Apr 11]; Available from: https://www.ncbi.nlm.nih.gov

10. Adetifa IMO, Uyoga S, Gitonga JN, Mugo D, Otiende M, Nyagwange J, et al. Temporal trends of SARS-CoV-2 seroprevalence during the first wave of the COVID-19 epidemic in Kenya. Nature Communications 2021 12:1 [Internet]. 2021 Jun 25 [cited 2023 Apr 13];12(1):1–6. Available from: https://www.nature.com/articles/s41467-021-24062-3

11. Kagucia EW, Gitonga JN, Kalu C, Ochomo E, Ochieng B, Kuya N, et al. Anti-Severe Acute Respiratory Syndrome Coronavirus 2 Immunoglobulin G Antibody Seroprevalence Among Truck Drivers and Assistants in Kenya. Open Forum Infect Dis [Internet]. 2021 Jul 1 [cited 2023 Apr 13];8(7). Available from: https://pubmed.ncbi.nlm.nih.gov/34660838/

12. Etyang AO, Lucinde R, Karanja H, Kalu C, Mugo D, Nyagwange J, et al. Seroprevalence of Antibodies to Severe Acute Respiratory Syndrome Coronavirus 2 Among Healthcare Workers in Kenya. Clin Infect Dis [Internet]. 2022 Jan 15 [cited 2023 Apr 13];74(2):288–93. Available from: https://pubmed.ncbi.nlm.nih.gov/33893491/

13. United Kingdom Health Surveillance data dashboard. Available from: https://ukhsa-dashboard.data.gov.uk/

