## Supplementary material for "Leveraging an Online Dashboard to Inform on Infectious Disease Surveillance: A case Study of COVID-19 in Kenya": Supplimental Appendix

### Appendix 1: Warning Signal for Potentially Growing Lineages of SARS-CoV-2 in Kenya

The GISAID database holds reported data on circulating variants of SARS-CoV-2 worldwide. This data can help track the spread and growth of potential variants of concern. We propose a mechanism to analyse the growth of variants in selected countries or regions and their potential to enter and grow in Kenya (Figure 1).

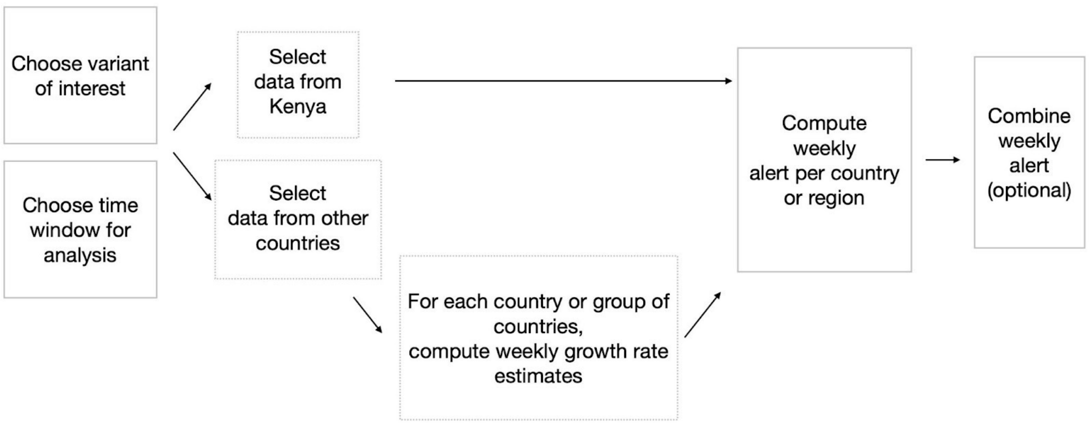


Figure 1. The process to estimate the growth of a variant in different countries or regions to produce a weekly warning signal.

First, a range of potential variants of concern is chosen for the analysis, based on emergent and newly identified sequences in Kenya. Data on reported sequences of the selected variants are extracted from GISAID and grouped by different countries, larger regions or continents. Then, the weekly growth rates of each variant and country are estimated using a Bayesian hierarchical model, which provides the posterior distribution of growth rates (Guzman-Rincon et al., 2023). A variant is labelled as "growing" if the probability of a positive growth rate is greater than 75%. Information on growing variants combined with their abundance in Kenya defines the strength of the alert, which can be labelled as "no evidence of growth", "evidence of slow growth" and "evidence of growing" (Figure 2).

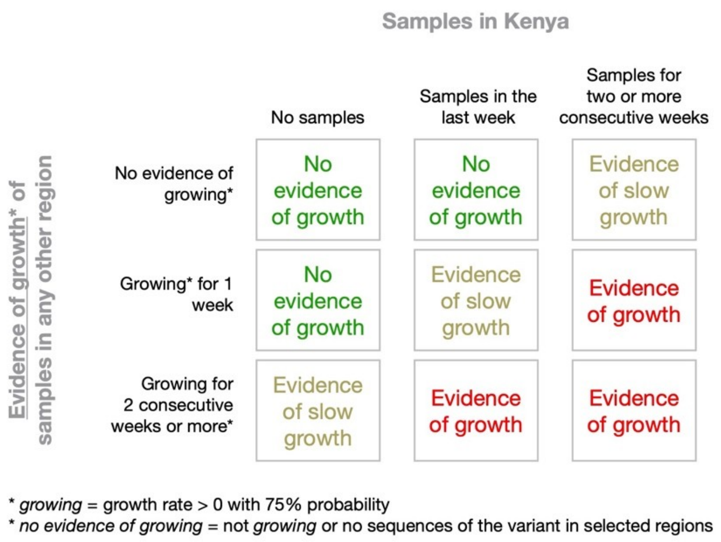


Figure 2. Strength of the alert for a given variant, based on the abundance of the variant in Kenya and the estimated growth in the country or region.

If multiple countries or regions are analysed, the alert can be combined. For instance, if a variant is identified with the "evidence of growing" alert in at least one country, the combined alert can take the same value.

Supplementary Table 1: Frequency of Data Update in the Dashboard.

| **Tab** | Section | Update Period | Comments |
|---|---|---|---|
| Home | Case Updates | Weekly | |
| | Testing Updates | UDA | |
| | Counties with Recent Updates | Weekly | |
| | Most Affected Counties | UDA | |
| | Trends in cases | Weekly | |
| | Trends in Fatalities | Weekly | |
| | Cases by age and gender | UDA | |
| | Fatalities by age and gender | UDA | |
| Counties | All the sections | UDA | |
| Vaccination | All the sections | Monthly | |
| Seroprevalence | All the sections | UDA | Depends on sero-study results |
| Variants | All the sections | Weekly | |
| ASS | All the sections | UDA | Depends on newly collected datasets |
| Phylogeny | All the sections | Monthly | |

UDA – Upon Data Availability.
